# Determinants of Periodic Health Examination Uptake: Insights from a Jordanian Cross-Sectional Study

**DOI:** 10.1101/2024.02.03.24302286

**Authors:** Abdul Aziz Tayoun

## Abstract

**Background:** Routine Periodic Health Examinations (PHE) for asymptomatic adults involve clinical preventive services, this routine process aims to prevent morbidity and mortality by identifying modifiable risk factors and early signs of treatable diseases.

PHE is a common procedure in primary healthcare worldwide, including Jordan. The country is undergoing an epidemiological transition toward non-communicable diseases (NCDs), which are the leading causes of morbidity and mortality The prevalence of smoking is among the highest in the world with escalating rates of obesity and physical inactivity. Notably, hypertension and diabetes mellitus are the most significant concerns.

**Objectives:** The goals of this study are to determine the extent to which individuals in Jordan participate in periodic health examinations and to evaluate the various sociodemographic, health-related, knowledge, and behavioural factors that influence this participation.

**Methods:** The research methodology employed in this study is a cross-sectional approach that includes 362 participants aged 18 years or older residing in Jordan. A convenient sampling method was utilized, and data were collected through an online questionnaire. The analysis involves the application of logistic regression through SPSS to investigate the relationship between various influencing factors and the uptake of periodic health examinations (PHE).

**Results:** Our study indicates that 27.1% of participants underwent periodic health examinations (PHE) within the last 2 years, with a 95% confidence interval ranging from 22.8% to 31.9%. Noteworthy predictors of PHE uptake among Jordanians include recent visits to a primary health care facility within the last year, monthly income, and knowledge levels regarding periodic health examinations and preventive health measures. These variables emerged as the strongest predictors in our analysis, shedding light on key factors influencing PHE participation in the population.

**Conclusion:** Periodic health examination uptake is notably low in Jordan. Key determinants of this uptake include recent visits to a primary health care facility within the last year, monthly income, and knowledge levels regarding periodic health examination (PHE) and preventive health services. To enhance PHE participation, there is a critical need for the integration of periodic health examinations with primary health care services in Jordan.

**WHAT IS ALREADY KNOWN ON THIS TOPIC:** In primary health care, periodic health examinations are a common procedure for asymptomatic individuals. These examinations are key in early disease detection and managing risk factors. However, the rate of uptake and the factors influencing this uptake in Jordan are not well-known.

**WHAT THIS STUDY ADDS:** This study provides, for the first time in Jordan, an estimation of the rate and predictive factors for the uptake of periodic health examinations.

**HOW THIS STUDY MIGHT AFFECT RESEARCH, PRACTICE OR POLICY:** The findings indicate a low rate of periodic health examination uptake in Jordan. The study suggests that integrating free periodic health examinations into primary health care services might increase awareness and uptake of these examinations. This has potential implications for health policy and practice, possibly leading to improved health outcomes through earlier disease detection and management.

## Introduction

Routine Periodic Health Examinations (PHE) for asymptomatic adults are integral components of primary healthcare. These examinations involve clinical preventive services administered by primary health care clinicians to individuals without evident signs or symptoms of illness, constituting a routine health care process. The overarching goal of these examinations is to proactively prevent morbidity and mortality. This is achieved by identifying modifiable risk factors and detecting early signs of treatable diseases(1).

The Health Belief Model (HBM) was conceptualized to elucidate the reasons behind individuals’ reluctance to engage in disease prevention programs and health check-ups. Serving as a crucial predictive framework, the HBM aids in understanding various health-related behaviours, spanning activities such as smoking, exercise, patient roles, and utilization of medical services(2).

Health beliefs, integral to the HBM, are defined as personal convictions associated with perceiving and managing specific diseases. These beliefs encompass key elements: perceived sensitivity, perceived severity, perceived benefit, perceived barrier, and cue to action(3).

In a systematic review recently published in the Canadian Family Physician journal, the primary objective was to assess the reasons for visit (RFV) to primary health care clinics. Notably, clinicians participating in the review identified routine health maintenance as the third most prevalent reason for individuals seeking consultations with primary health care physicians. This ranking positioned routine health maintenance after upper respiratory tract infections and hypertension, highlighting its significant role in motivating individuals to engage with primary health care services(4).

In a study conducted among undergraduate students in a Nigerian health science college, it was found that a substantial 91.2% of participants demonstrated awareness of periodic health examinations (PHE). However, the actual participation in PHE was notably low at 28.4%. The primary hindrance to uptake was identified as insufficient time. Additional barriers encompassed factors such as religious considerations, duration of education, perceived susceptibility to diseases, financial constraints, apprehension about the results, and a general lack of interest(5).

A nationwide study conducted in Saudi Arabia revealed that 22.9% of participants aged 15 years or older had undergone a Periodic Health Examination (PHE) in the preceding 2 years. The probability of receiving a PHE during this period exhibited positive correlations with various factors, including age, educational attainment, marital status, regular consumption of five servings of fruits and vegetables daily, and the presence of diagnoses such as prediabetes, diabetes, or hypercholesterolemia. Furthermore, individuals who had visited a healthcare setting within the last 2 years due to illness or injury were also more likely to have undergone a PHE(6).

Jordan, classified as an upper-middle-income country, spans an area of 89,318 square kilometers, divided into 12 governorates. The population has undergone substantial growth, increasing from 5.4 million in 2003 to over 11.5 million in 2023. This demographic shift can be attributed largely to the influx of refugees and a relatively high birth rate(7,8).

The country is currently undergoing a notable epidemiological transition characterized by a rising prevalence of non-communicable diseases (NCDs). These diseases now constitute approximately 78% of deaths, establishing themselves as the primary cause of mortality and morbidity among the Jordanian population. Key risk factors contributing to the burden of NCDs include tobacco use, with a prevalence of about 50% (including e-cigarettes and shisha). One-quarter of the population reports insufficient physical activity, and approximately 60% are classified as overweight or obese. Additionally, 22% of the population is hypertensive, 14% is diabetic, and the prevalence of depression is about 18% (9).

The presented profile underscores a pressing concern regarding the high risk of Non-Communicable Diseases (NCDs) in the country. Evidently, there is a need for evidence-based preventive health measures to curb the progression of NCDs and their associated risk factors. Periodic health examinations, if conducted in accordance with evidence-based guidelines, have the potential to serve as an effective tool in controlling both communicable and NCDs.

Recognizing the urgency of the situation, it is imperative to gather data on the uptake rate of periodic health examinations and identify the factors influencing this uptake. The absence of previous studies on the uptake of periodic health examinations in Jordan underscores the necessity for comprehensive research. Our study aims to estimate the uptake of periodic health examinations among Jordanians, while concurrently investigating various sociodemographic, health status, knowledge, and behavioral factors that play a role in influencing this uptake. The findings from this research will not only contribute valuable insights into the current scenario but will also guide educational and promotional activities to encourage citizens to utilize preventive health services. In doing so, we strive to fill a crucial gap in existing knowledge and provide a foundation for evidence-based strategies to enhance public health in the country.

## Methodology

### Design

This descriptive cross-sectional study aims to estimate the uptake of Periodic Health Examination (PHE) and understand the various factors influencing this uptake. The research employs a questionnaire designed with five key domains: sociodemographic information, health status, PHE uptake history, knowledge about periodic health examination, and health behaviours based on the Health Belief Model (HBM). This comprehensive questionnaire allows for a thorough exploration of the factors impacting PHE utilization.

To ensure the representativeness of the sample and to minimize selection bias, the study adopts a proportional sampling strategy across four provinces of Jordan. This approach is carefully extended to maintain balance in gender and nationality among participants, thereby enhancing the study’s generalizability to the wider Jordanian population.

Acknowledging the challenges inherent in online research, particularly concerning measurement bias, the study incorporates several strategies to enhance data quality and participant understanding. The initial page of the online questionnaire explicitly outlines the study’s objectives and provides detailed instructions on how to complete the questionnaire. This effort is complemented by the availability of the researcher for any clarifications, ensuring participants’ queries or doubts can be promptly addressed.

### Target population and sampling

The survey was conducted on March 15, 2023, and respondents were recruited online via Google Forms. The target population for this survey comprises Jordanian citizens who are 18 years of age or older.

*Inclusion Criteria: Any* citizen, regardless of nationality, aged 18 years or above and residing in Jordan.

Exclusion Criteria: Persons below the age of 18, and individuals who decline to participate in the study.

We recruited a total of 362 respondents, aiming to provide a representative sample that reflects the entire population of Jordan in terms of district, age, sex, and nationality. The convenient sample size of 362 was calculated using the sample size formula for proportions:

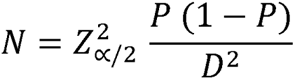

This calculation considered a study conducted in Saudi Arabia where approximately 34% of the population underwent periodic health examinations(10). The chosen values for statistical significance (α error) and margin of error (D) were 0.05 and 5%, respectively. As a result, the calculated sample size required for the survey was determined to be 345 respondents.

### Questionnaire Development

The PHE questionnaire, comprising 36 questions across five domains, was developed following an extensive literature review (10–14). The questionnaire’s structure is as follows:

- Sociodemographic Domain (9 items): Inquires about relevant sociodemographic variables of participants.
- Health Status and Risk Factors Domain (7 items): Explores participants’ health status and associated risk factors.
- PHE Uptake Domain (4 questions): Focuses on the outcome variable of periodic health examination.
- Knowledge about PHE and Preventive Health Services Domain (8 items): Assesses knowledge using a 3-option scale (’agree,’ ’don’t agree,’ ’I don’t know’). The items are scored, with correct answers receiving a score of 1 and incorrect or ’I don’t know’ responses scored as 0. The total score ranges from 0 to 8, with higher scores indicating greater knowledge of health check-ups and preventive measures. Cronbach’s α, estimated during the pilot phase with 25 participants, was found to be 0.682.
- Health Behaviors towards PHE based on HBM Domain (6 items): Measures health behaviors using a 5-point Likert scale ranging from 1 (’strongly disagree’) to 5 (’strongly agree’). The total scores range from 6 to 30, with higher scores indicating more positive health beliefs for each item. The Cronbach alpha for health behaviors towards PHE during the pilot testing phase was found to be 0.736, demonstrating acceptable internal consistency.

The questionnaire was translated into Arabic for comprehensibility and then translated back to English with the assistance of an expert translator. This rigorous process ensures the clarity and accuracy of the questionnaire across languages.

### Statistical analysis

The main outcome variable is the uptake of periodic health examinations in Jordan, categorized as a dichotomous (yes or no) variable. The independent variables encompass sociodemographic, health status, knowledge, and health behavioural factors. To ensure the integrity of the analysis, records with missing data were excluded. Data analysis was conducted using IBM SPSS, version 27.0 (IBM, Armonk, NY, USA).

Participant characteristics were examined using counts, percentages, means, and standard deviations (SD) through descriptive statistics. Graphs and tables were employed as needed for visual representation. A 95% confidence interval was calculated using appropriate methods, and a 2-sided p-value less than 0.05 was considered statistically significant.

Associations between continuous variables and the outcome variable were assessed using the independent student-t test, while the Chi-square independent test measured associations between categorical variables and the outcome variable. Multivariate logistic regression analysis was employed to examine the relationship between the uptake of periodic health examination and various independent co-variables.

Additionally, a hierarchical block-wise logistic regression model was constructed to identify the most powerful predictor variables. This comprehensive approach blends descriptive, inferential, and multivariate statistical techniques to provide a thorough understanding of the factors influencing the uptake of periodic health examination in Jordan.

### Ethical considerations

Participants must be 18 years of age or older to be eligible for the study. Data collection was conducted through Google Forms, and participants could only proceed by clicking the "agree" button; the survey couldn’t be completed if they chose "don’t agree." The first web page of the online survey outlined guidelines, including the study’s objectives, procedures, and data collection methods.

All collected data are treated with strict confidentiality—no personal identifiers such as ID, email, or other information are required. Data is securely stored on a password-protected computer, accessible only to researchers and authorised personnel. Participant withdrawal from the study ensures their responses will not be included in data analysis, and no disadvantages are incurred by terminating or rejecting the survey.

Contact information for the researcher was provided for clarification purposes. The research protocol underwent ethical review and received approval from the Research Ethics Committee of Jordan University (IRB No. 13-2023) and the Jordan University Hospital Ethical Committee (IRB No. 10/2023/4560).

## Results

Between March and April 2023, 365 individuals participated in the study. Three participants were excluded (one was less than 18 years old, and the other two did not complete the questionnaire), leaving 362 participants for analysis

### Descriptive statistics

The demographic characteristics of participants are summarized in Table 1. The mean age was 38.22 years (range 18 to 88 years, SD 14.63), with slightly more males (51.1%). Approximately 63% were married, 74.6% were Jordanians, and 55.8% held a university degree. Figure 1 indicates that a majority (62.2%) reported a monthly income of less than 500 Jordan dinars (700 USD), with half lacking health insurance.

**Figure 1.**
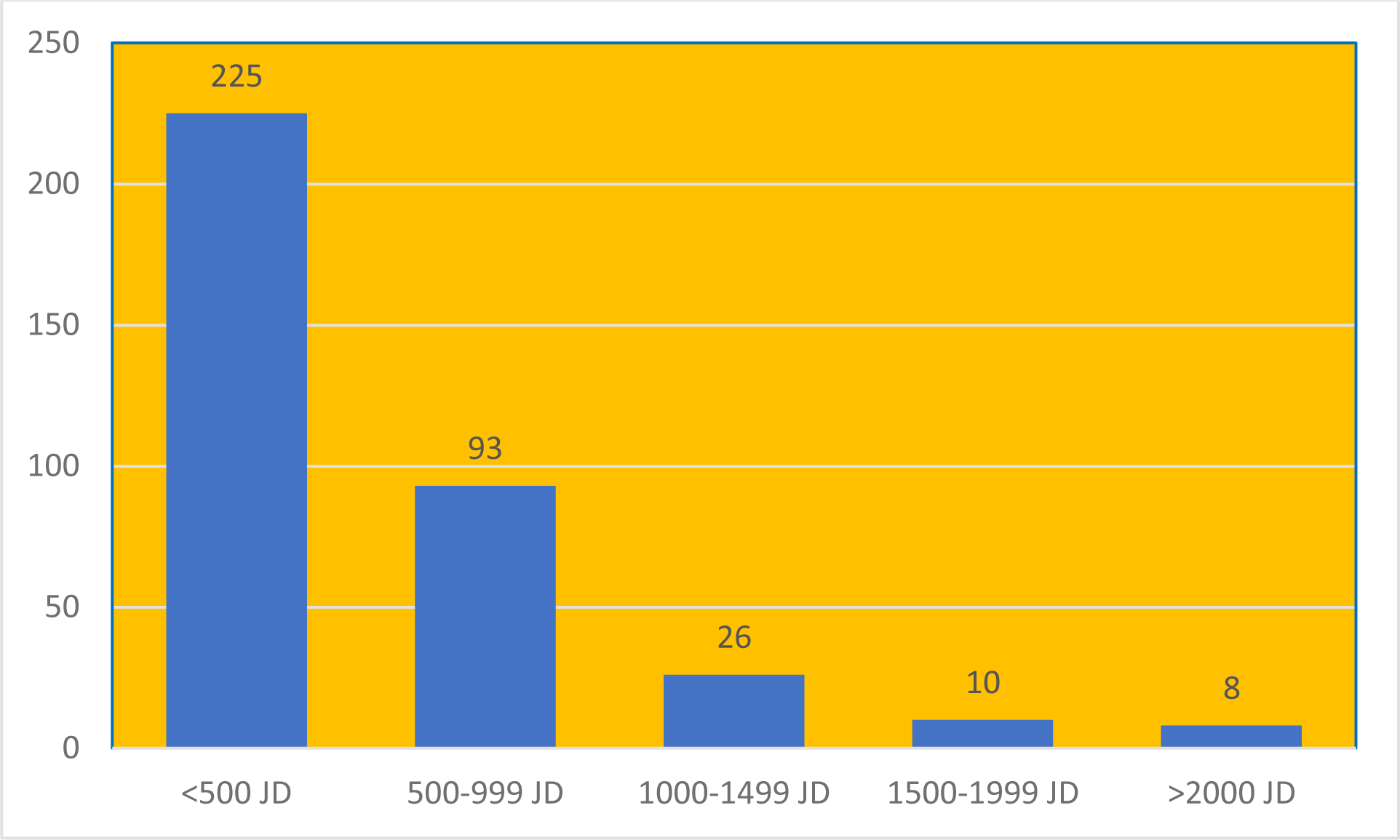
Monthly income of PHE study participants in Jordan, 2023. PHE : periodic health examination.

**Table 1.**
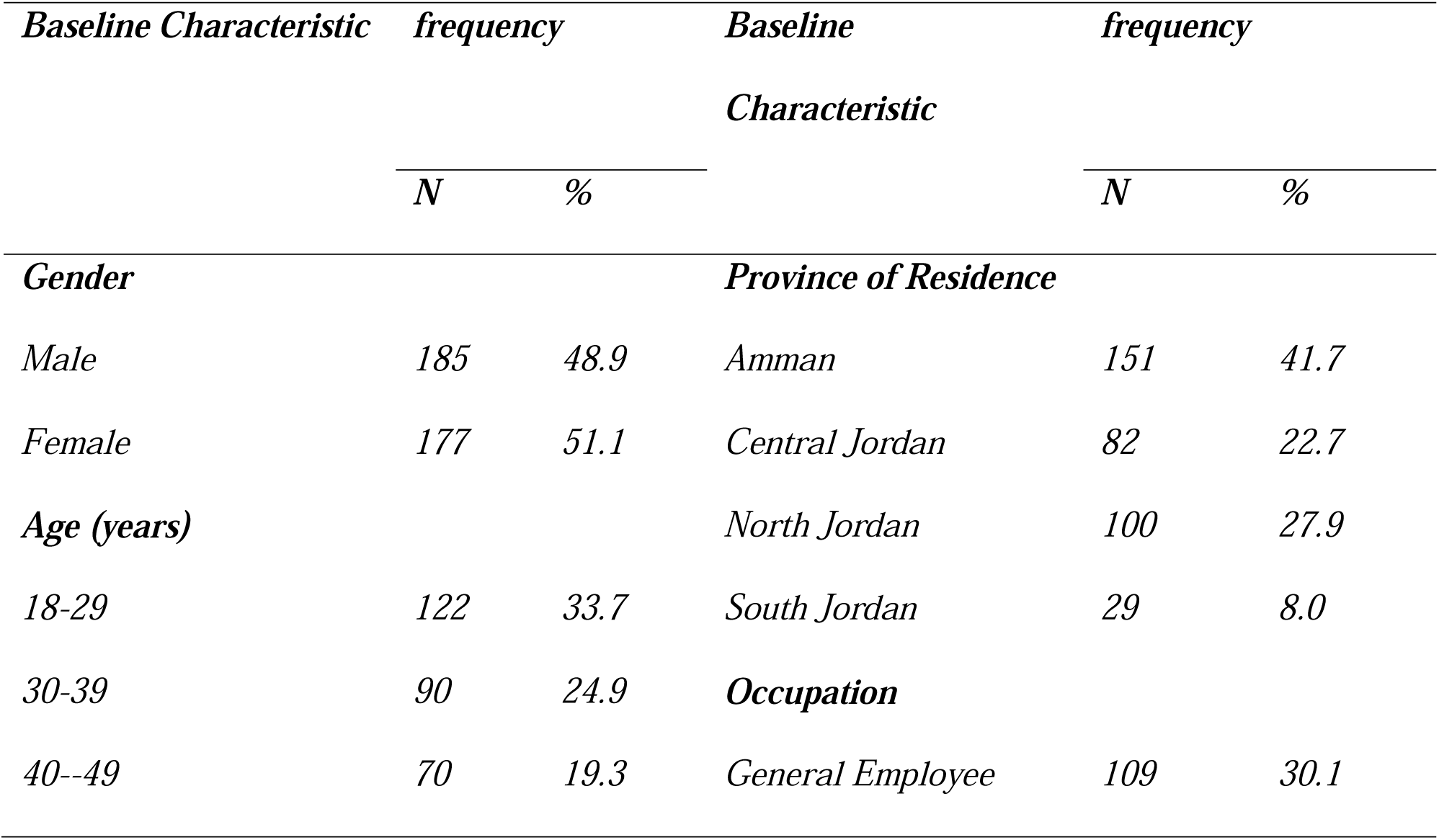
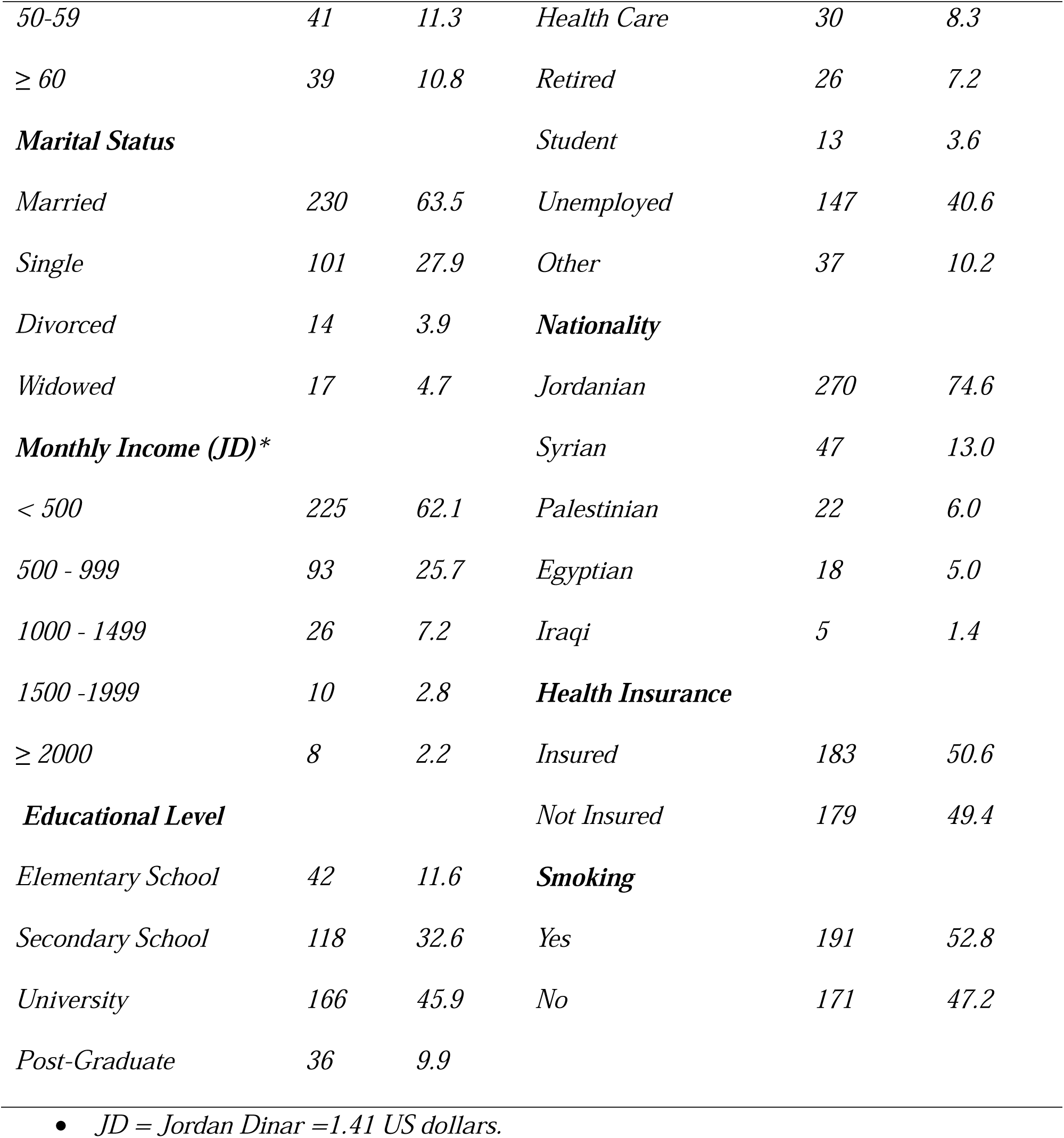
Sociodemographic Characteristics of Participants (N=362) in the study “Periodic health examination in Jordan “,2023.

| ***Baseline Characteristic*** | ***frequency*** |  | ***Baseline Characteristic*** | ***frequency*** |  |
| --- | --- | --- | --- | --- | --- |
|  | ***N*** | ***%*** |  | ***N*** | ***%*** |
| ***Gender*** |  |  | ***Province of Residence*** |  |  |
| *Male* | 185 | 48.9 | *Amman* | 151 | 41.7 |
| *Female* | 177 | 51.1 | *Central Jordan* | 82 | 22.7 |
| ***Age (years)*** |  |  | *North Jordan* | 100 | 27.9 |
| *18-29* | 122 | 33.7 | *South Jordan* | 29 | 8.0 |
| *30-39* | 90 | 24.9 | ***Occupation*** |  |  |
| *40--49* | 70 | 19.3 | *General Employee* | 109 | 30.1 |
| 50-59 | 41 | 11.3 | Health Care | 30 | 8.3 |
| ≥ 60 | 39 | 10.8 | Retired | 26 | 7.2 |
| **Marital Status** |  |  | Student | 13 | 3.6 |
| Married | 230 | 63.5 | Unemployed | 147 | 40.6 |
| Single | 101 | 27.9 | Other | 37 | 10.2 |
| Divorced | 14 | 3.9 | **Nationality** |  |  |
| Widowed | 17 | 4.7 | Jordanian | 270 | 74.6 |
| **Monthly Income (JD)\*** |  |  | Syrian | 47 | 13.0 |
| < 500 | 225 | 62.1 | Palestinian | 22 | 6.0 |
| 500 - 999 | 93 | 25.7 | Egyptian | 18 | 5.0 |
| 1000 - 1499 | 26 | 7.2 | Iraqi | 5 | 1.4 |
| 1500 -1999 | 10 | 2.8 | **Health Insurance** |  |  |
| ≥ 2000 | 8 | 2.2 | Insured | 183 | 50.6 |
| **Educational Level** |  |  | Not Insured | 179 | 49.4 |
| Elementary School | 42 | 11.6 | **Smoking** |  |  |
| Secondary School | 118 | 32.6 | Yes | 191 | 52.8 |
| University | 166 | 45.9 | No | 171 | 47.2 |
| Post-Graduate | 36 | 9.9 |  |  |  |

- - JD = Jordan Dinar =1.41 US dollars.

Regarding health status, 66.3% reported very good or excellent health, 21.5% had a chronic disease, and 55% visited a primary healthcare clinic in the past year as clearly seen in Figure 2 . Additionally, 52.8% were current smokers.

**Figure 2:**
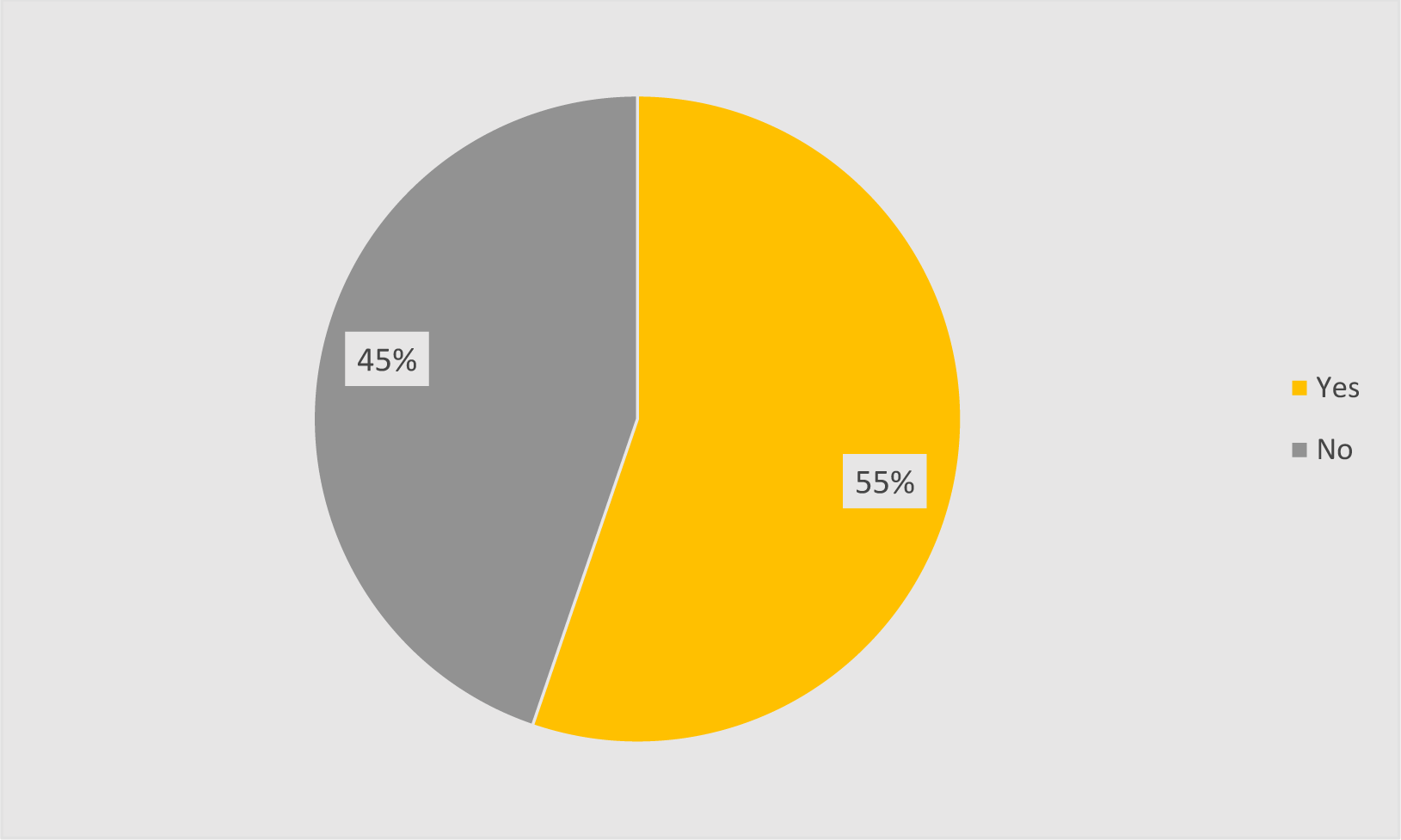
Visit to primary health care facility of participants of PHE uptake study, Jordan 2023.

Concerning periodic health examination (PHE), 27.1% (95% CI 22.8 to 31.9) of participants underwent a medical check-up in the last 2 years.

### Logistic regression analysis

When analyzing the predictor factors associated with the uptake of Periodic Health Examination (PHE), the Forest plot in Figure 3 highlights several significant findings:

**Figure 3:**
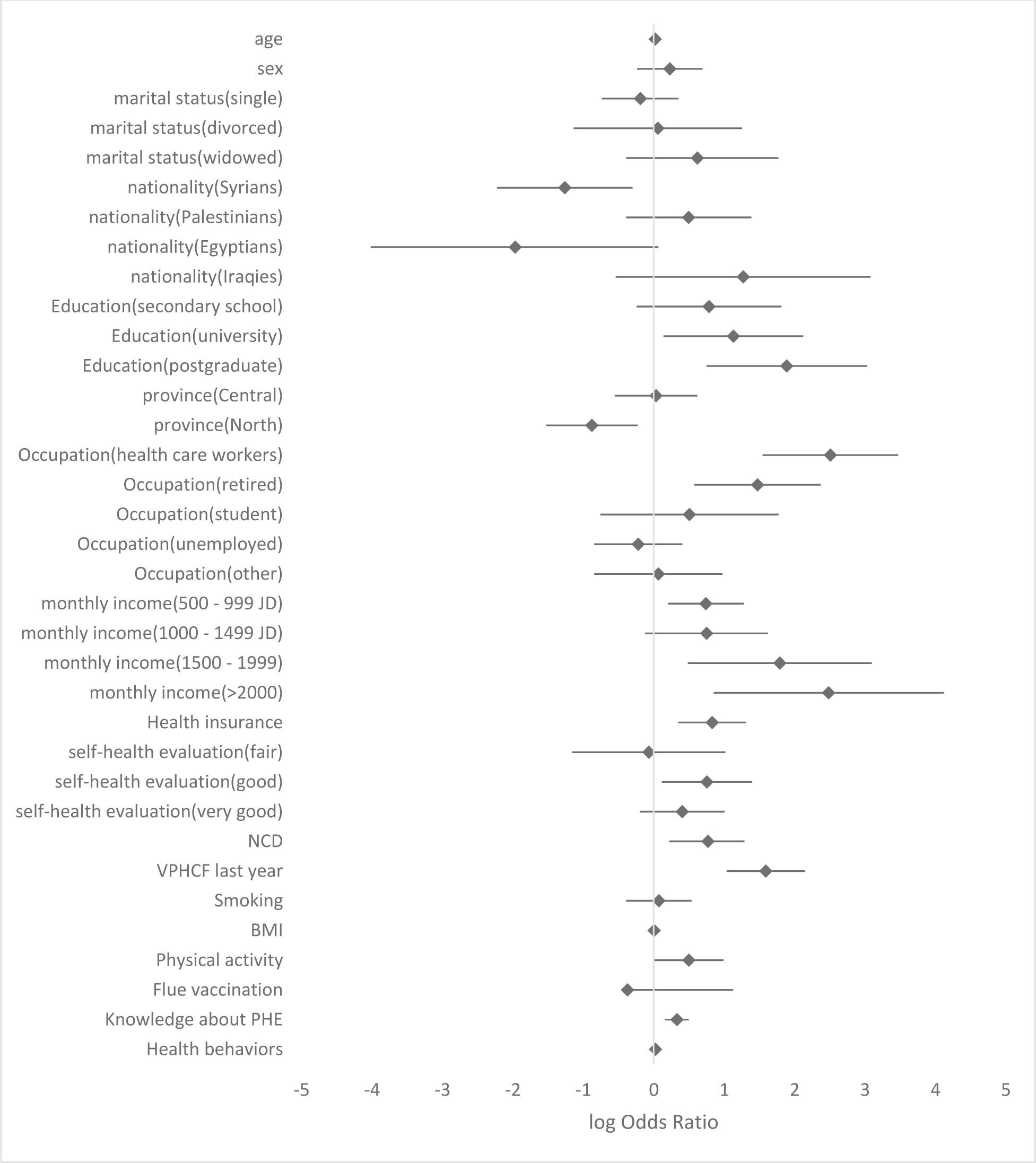
**univariate logistic regression analysis for predictor factors of PHE uptake, Jordan 2023.** PHE: periodic health examination. JD: Jordanian dinar. VPHCF: visit to primary health care facility BMI : body mass index

**Age** played a significant role, revealing that with each additional year, the odds of undertaking PHE increased by 2.2% (P-value 0.006, CI 1.006 to 1.038). **Nationality** also proved to be a factor, with Syrians demonstrating a lower frequency of PHE uptake, the odds of Syrians undergoing PHE were 0.283 times compared to Jordanians (P-value= 0.010, 95% CI 0.108 to 0.741). **Education level** exhibited a strong association, with postgraduates displaying 6.62 times higher odds of undertaking PHE compared to individuals with only primary school education (P-value= 0.001, CI 2.117 to 20.709). O**ccupation**, specifically within the healthcare sector, showed a significant impact. Health care workers displayed odds 12.286 times higher for undergoing PHE compared to general employees (P-value 0.000, CI 4.69 to 32.19). **Monthly income** levels were another influential factor, with individuals earning more than 2000 JD exhibiting 12.00 times the odds of receiving PHE compared to those with a monthly income of less than 500 JD (P-value= 0.003, CI 2.343 to 61.449). **Health insurance** emerged as a significant facilitator of PHE uptake. Insured participants demonstrated odds 2.3 times higher for undertaking PHE compared to non-insured individuals (P-value=0.001, CI 1.417 to 3.712).

Regarding health factors, Individuals with a chr**onic disease** have 2.163 times the odds of undertaking PHE compared to persons without a chronic disease (p-value = 0.005, CI 1.258 to 3.629).Those who **visited a primary health care clinic** in the past year exhibit a significant impact on PHE uptake. They have 5 times the odds of PHE uptake compared to those who did not visit a Primary Health Care Facility (PHCF) in the past year (p-value = 0.000, CI2.817to8.570). Individuals with sufficient **physical activity** have 1.649 times the odds of undertaking PHE than those without enough physical activity (p-value = 0.046, CI 1.009 to 2.693). And finally for every extra point in **health knowledge** about periodic health examination, there is a notable 39% increase in PHE uptake (p-value = 0.000, CI 1.175 to 1.645).

On the other hand, several variables were **not found to be associated** with periodic health examination uptake. These included gender (P-value = 0.334), smoking status (P-value = 0.759), marital status (P-value = 0.524), health status self-evaluation (P-value = 0.184), seasonal influenza vaccination (P-value = 0.069), health behavior factors combined (P-value = 0.340), and BMI (P-value = 0.758).

### Adjusted Logistic Regression Model

After meticulous adjustment for confounding variables and the careful selection of both clinically and statistically significant factors, we successfully constructed a logistic regression model using the hierarchical block-wise method . This refined model, depicted in Table 2 encapsulates three variables that significantly influence the uptake of Periodic Health Examination (PHE).

**Table 2:**
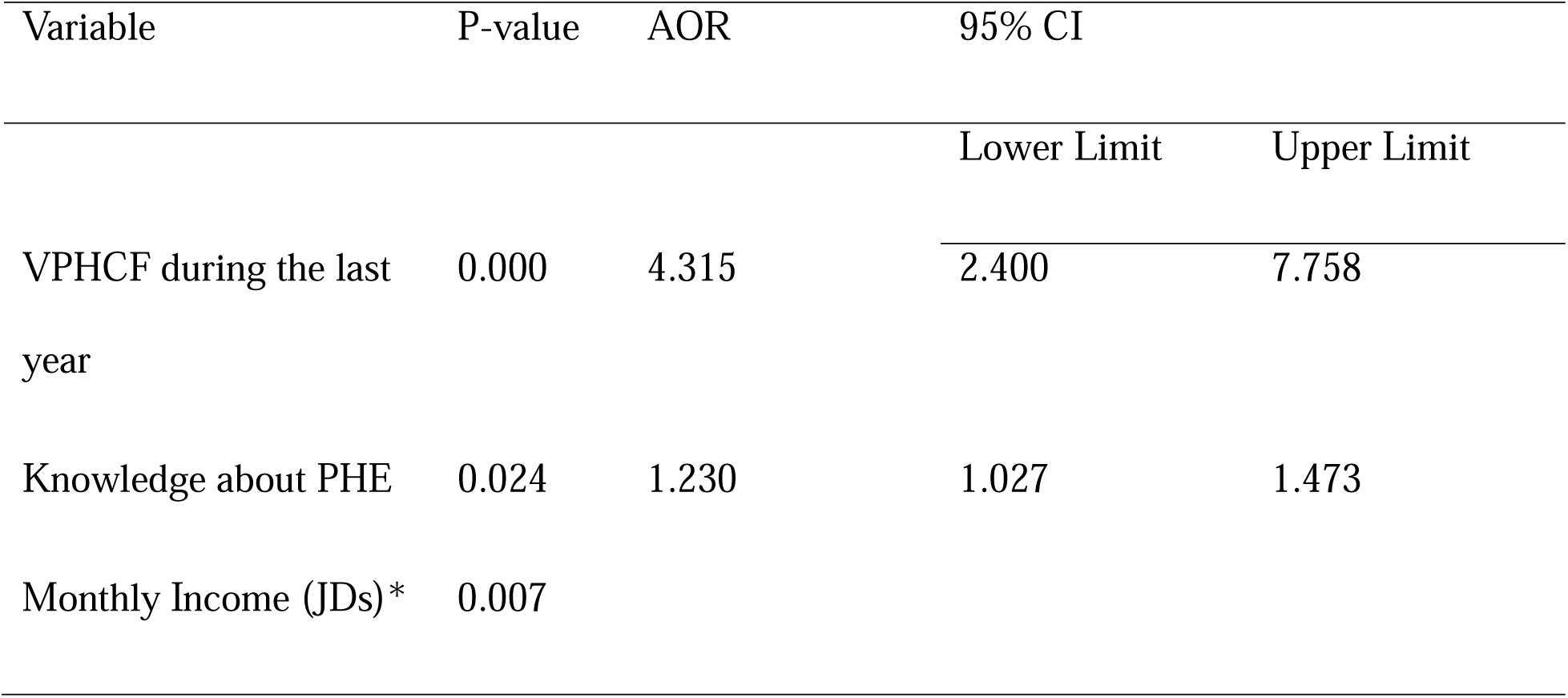
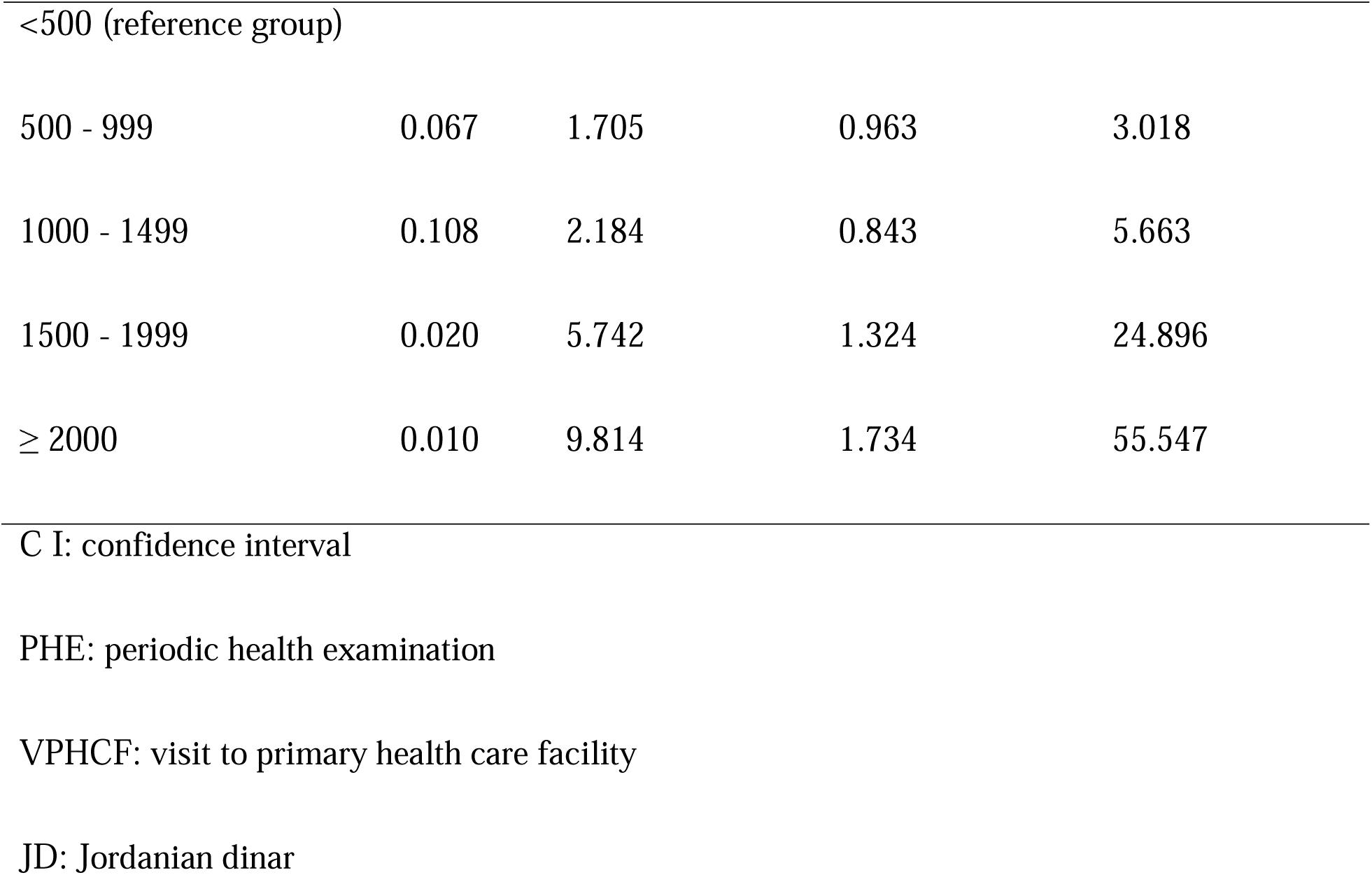
Logistic regression model for most significant predictor factors for PHE uptake, Jordan.

| Variable | P-value | AOR | 95% CI |  |
| --- | --- | --- | --- | --- |
|  |  |  | Lower Limit | Upper Limit |
| VPHCF during the last year | 0.000 | 4.315 | 2.400 | 7.758 |
| Knowledge about PHE | 0.024 | 1.230 | 1.027 | 1.473 |
| Monthly Income (JDs)* | 0.007 |  |  |  |
| <500 (reference group) |  |  |  |  |
| 500 - 999 | 0.067 | 1.705 | 0.963 | 3.018 |
| 1000 - 1499 | 0.108 | 2.184 | 0.843 | 5.663 |
| 1500 - 1999 | 0.020 | 5.742 | 1.324 | 24.896 |
| ≥ 2000 | 0.010 | 9.814 | 1.734 | 55.547 |

C I: confidence interval
PHE: periodic health examination
VPHCF: visit to primary health care facility
JD: Jordanian dinar

#### · Visit to PHCF in the Past Year

Individuals who visited Primary Health Care Facilities (PHCF) within the past year exhibited a substantial impact on PHE uptake. They demonstrated 4.315 times the odds of undertaking PHE compared to those who did not visit PHCF within the same timeframe (p-value = 0.000, 95% CI 2.400 to 7.758).

#### · Income Levels

Individuals with a monthly income of 1500-2000 JD displayed 5.742 times the odds of undertaking PHE compared to those with a monthly income of less than 500 JD (p-value = 0.020, 95% CI 1.324 to 24.896). Furthermore, those with a monthly income of more than 2000 JD exhibited even higher odds, with a value of 9.814 (p-value = 0.020, CI 1.734 to 55.547).

#### · Health Knowledge

The analysis indicates that for each point increase in PHE knowledge, there is a notable 23% increase in the likelihood of individuals opting for periodic health examinations (p-value = 0.024, CI 1.027 to 1.473).

## Discussion

Our study represents a pioneering effort as the first of its kind to assess the uptake of Periodic Health Examinations (PHE) and explore the various factors influencing this uptake specifically in Jordan. Our findings reveal that approximately 27.1% of Jordanians underwent PHE within the last 2 years, with a 95% confidence interval ranging from 22.8% to 31.9%.

Interestingly, our observed uptake rate is comparable to findings from studies conducted in Saudi Arabia (6,10) and Nigeria (12). However, it notably falls below the rates reported in studies conducted in the United States (1), the United Kingdom (13), and Switzerland (15).

These comparative statistics highlight variations in PHE utilization across different regions, suggesting potential differences in healthcare practices, accessibility, or public awareness. The similarities with Saudi Arabia and Nigeria could indicate shared regional trends, while the disparities with the USA, UK, and Switzerland may stem from distinct healthcare systems, cultural influences, or healthcare policies.

In our study, we identified the most influential determinant of Periodic Health Examination (PHE) uptake as a **recent visit to a primary health care facility (PHCF) within the last year**

This finding aligns with consistent results observed in several studies (6,16,17). Notably, individuals who had visited a PHCF in the last year were found to be five times more likely to undergo PHE compared to those who did not have such recent visits. This significant association persisted even after adjusting for other relevant factors, underlining the robustness of this relationship.

The second critical factor influencing PHE uptake in our study was **monthly income**. This result is in concordance with findings from various studies (1,12,14,17–21). The impact of monthly income on PHE uptake underscores the role of socio-economic factors in shaping healthcare-seeking behaviors, emphasizing the need for targeted interventions to address disparities.

The third influential factor identified in our study was **knowledge about PHE**. This result is consistent with previous research (22–24) and highlights the importance of informed decision-making in healthcare utilization. It is worth noting that knowledge about PHE is intertwined with other factors such as educational level and occupation. However, even after adjustment for these related factors, the association between knowledge about PHE and uptake remained robust.

Several additional variables were identified as associated with the uptake of Periodic Health Examination (PHE), providing a nuanced understanding of factors influencing preventive healthcare practices.

**Age**: The findings revealed a positive association between age and PHE uptake, aligning with results from other studies (13,17,19). This suggests that as individuals age, they are more likely to engage in routine health examinations, possibly due to increased awareness of health-related issues or a greater emphasis on preventive care with advancing age.

### Nationality

Notably, individuals of Syrian nationality were found to be less likely to undergo PHE compared to Jordanians. Economic factors may contribute to this difference, emphasizing the need for targeted interventions to ensure equitable access to preventive healthcare services among diverse populations.

### Place of Residence

Individuals residing in the North region were less likely to undergo PHE compared to those living in Amman city. This regional disparity suggests potential variations in healthcare accessibility or awareness, pointing towards the importance of tailoring interventions to specific geographic locations.

### Educational Level

Educational attainment exhibited a strong association with PHE uptake, with a noticeable increase in uptake corresponding to higher levels of education. This finding is consistent with results from other studies (17,21,25).

### Occupation

Health care workers and retired individuals were found to be more likely to undergo PHE compared to employees in general. The higher likelihood of PHE uptake among healthcare workers may be attributed to a heightened awareness of the importance of preventive healthcare within this group. For retired individuals, age may act as a confounding factor, influencing both retirement status and PHE uptake.

Our study identified several **health-related factors** associated with the uptake of Periodic Health Examination (PHE). These factors include the presence of **chronic diseases** supported by other studies (6,14,18,22,26), **being insured** supported by studies (17,21,25,27,28), and engagement in **physical activity**(1), a correlation consistent with numerous studies. Understanding these health-related determinants provides valuable insights for healthcare providers and policymakers seeking to enhance preventive healthcare practices.

Our study found that certain factors **were not significantly associated** with the uptake of Periodic Health Examination (PHE). These include sex, contrary to numerous studies that found females more likely to undertake PHE than males (6,13,15,20). Marital status, which is often linked to higher PHE uptake in previous studies (1,13–15,19,29,30), did not show a significant association in our research. Surprisingly, smoking status was not found to be associated with the uptake of PHE, contrary to findings from several studies that suggested smokers are less likely to undergo PHE than non-smokers (11,13,15,20,29). These non-associated factors highlight the complexity of health behavior determinants and suggest variations within the specific context of the studied population.

Our study found no significant association between combined behavioral factors and the uptake of Periodic Health Examination (PHE), contradicting findings in many studies (3,11,14,20,31,32). Possible explanations include questionnaire suitability for the Jordanian population or participant comprehension issues, emphasizing the need for culturally sensitive research methodologies and clear survey instruments in future studies.

#### Limitations of the Study

- Cross-Sectional Design: The study’s cross-sectional nature limits its ability to establish causal relationships between independent and outcome variable .
- Convenience Sampling: Utilizing a convenience sample may lead to selection bias, affecting the representativeness of the results.
- Self-Administered Online Questionnaire: The reliance on self-reported data through an online questionnaire could introduce measurement bias, as it depends on participants’ honesty and self-perception.

#### Strengths of the Study

- First of Its Kind: This study is pioneering in examining the prevalence and determinants of Periodic Health Examination (PHE) across Jordan, offering new insights into a previously under-researched area.
- Broad Geographic Coverage: It encompasses the entire population across Jordan’s four provinces, enhancing the study’s representativeness and applicability to the Jordanian population.
- Proportional Representation: The sample includes participants proportional to gender and nationality distribution throughout the country, contributing to the generalizability of the findings.
- Statistical Methods to Address Bias: To mitigate confounding bias, the study employs adjusted odds ratios and hierarchical block-wise logistic regression, constructing a comprehensive model for analysis.

#### Implications for Clinical Practice

- Integration of Preventive and Primary Health Care Services: The most significant predictor of Periodic Health Examination (PHE) uptake is a recent visit to Primary Health Care Facility (VPHCF). We recommend integrating preventive health services with primary healthcare to enhance accessibility and efficiency(33), providing incentives for both healthcare providers and patients(34).
- Offering free preventive services: To address the impact of economic factors on PHE uptake, we propose integrating free preventive services within the primary healthcare context. Negotiations with private health insurance companies to include preventive services, including PHE, in their coverage can further promote accessibility(34).
- Increasing Awareness through Campaigns: The positive association between knowledge about PHE and uptake suggests a need for increased awareness. Organized and evidence-based campaigns can be effective in raising awareness about PHE and preventive measures in general.
- Guideline Development and Implementation: To enhance preventive care services, we recommend developing and implementing guidelines tailored to age- and sex-specific examinations. Drawing inspiration from successful models like the Canadian Task Force on PHE(35) and the US Taskforce for Preventive Services can guide this process(36).
- Memory Aid for Primary Care Physicians: Providing primary care physicians with a memory aid based on evidence-based practices can improve the quality of care during PHE(37). Such aids can include lists of examinations tailored to patients’ age, sex, and health conditions.

Implementing these recommendations can strengthen Jordan’s preventive healthcare services, contributing to improved population health outcomes.

#### Implications for Future Research

- Exploring Behavioral Factors in Jordan: Our study did not find a relationship between behavioral factors (sensitivity, self-efficacy, perceived severity, and perceived benefit) and Periodic Health Examination (PHE) uptake. Given that other studies have identified such relationships, we recommend further investigation of these factors in the Jordanian context in future studies.
- In-Depth Examination of Smoking as a Predictor: Our study suggests that smoking was not associated with PHE uptake, contrary to findings in other studies. We recommend conducting more detailed and specific studies on smoking as a predictor factor for PHE uptake in the Jordanian population to better understand the nuances and contributing factors.

By addressing these areas in future research, a more comprehensive understanding of the determinants and predictors of PHE uptake in Jordan can be gained, contributing to the development of targeted interventions and healthcare policies.

## Conclusion

In conclusion, our study sheds light on the low uptake of periodic health examinations (PHE) in Jordan. The analysis identifies visitation to primary health care facilities in the past year, monthly income, and knowledge about PHE and preventive health services as crucial predictors influencing the likelihood of undergoing periodic health examinations.

The findings underscore the need for targeted interventions aimed at increasing awareness and knowledge about the importance of preventive health practices, particularly among individuals with lower income levels. Additionally, the observed association between regular visits to primary health care facilities and higher PHE uptake suggests an opportunity for integrating periodic health examinations with existing primary health care services.

## Data Availability

All data produced in the present study are available upon reasonable request to the author at

